# Disentangling Increased Testing from Covid-19 Epidemic Spread

**DOI:** 10.1101/2020.07.09.20141762

**Authors:** Benjamin Lengerich, Willie Neiswanger, Eugene J. Lengerich, Eric P. Xing

## Abstract

To design effective disease control strategies, it is critical to understand the incidence of diseases. In the Covid-19 epidemic in the United States (caused by outbreak of the SARS-CoV-2 virus), testing capacity was initially very limited and has been increasing at the same time as the virus has been spreading. When estimating the incidence, it can be difficult to distinguish whether increased numbers of positive tests stem from increases in the spread of the virus or increases in testing. This has made it very difficult to identify locations in which the epidemic poses the largest public health risks. Here, we use a probabilistic model to quantify beliefs about testing strategies and understand implications regarding incidence. We apply this model to estimate the incidence in each state of the United States, and find that: (1) the Covid-19 epidemic is likely to be more widespread than reported by limited testing, (2) the Covid-19 epidemic growth in the summer months is likely smaller than it was during the spring months, and (3) the regions which are at highest risk of Covid-19 epidemic outbreaks are not always those with the largest number of positive test results.

## Introduction

In mid-June 2020, the number of positive PCR test results for SARS-CoV-2 infection began to increase in selected US states for the first sustained period since the epidemic appeared to peak in early-April 2020. This apparent resurgence has led to calls for additional public health strategies to reduce risk of infection. However, there has also been public and scientific debate about the potential influence of increased testing on the apparent increase in the incidence of SARS-CoV-2 infection. This debate has led to the need to disentangle the increase in the incidence from the increase in the number of administered tests; however, few rigorous quantitative efforts have been reported. In this paper, we propose a probabilistic model to disentangle these factors and then apply the model to identify states experiencing increases in SARS-CoV-2 incidence. Using this model, we are able to better estimate the incidence of SARS-CoV-2, and rank the states in terms of their incidence after correcting for expanded testing.

Jupyter notebooks to perform these estimates and daily-updated results are available at https://github.com/blengerich/Covid19-latentcases.

## Results

### Estimated Incidence in States Over Time

Fig. 1 plots the estimated daily incidence 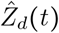 calculated according to Eq. (9) for six states of various population densities. Results for all 50 states are available on Github^1^. In each panel, we plot the state population, the daily number of positive tests *ñ*_*dp*_(*t*), the daily number of negative tests *ñ*_*dp*_(*t*), and the estimated daily incidence under different testing strategies quantified as *c*(*t*), the propensity-to-test infected individuals relative to uninfected individuals. In addition, we also plot the current number of patients hospitalized with Covid-19—this number tends to be greater than the daily number of positive SARS-CoV-2 tests because patients can remain hospitalized for several days, but the curve is discontinuous because states have changed the procedures for reporting this number over the course of the epidemic.

**Figure 1:**
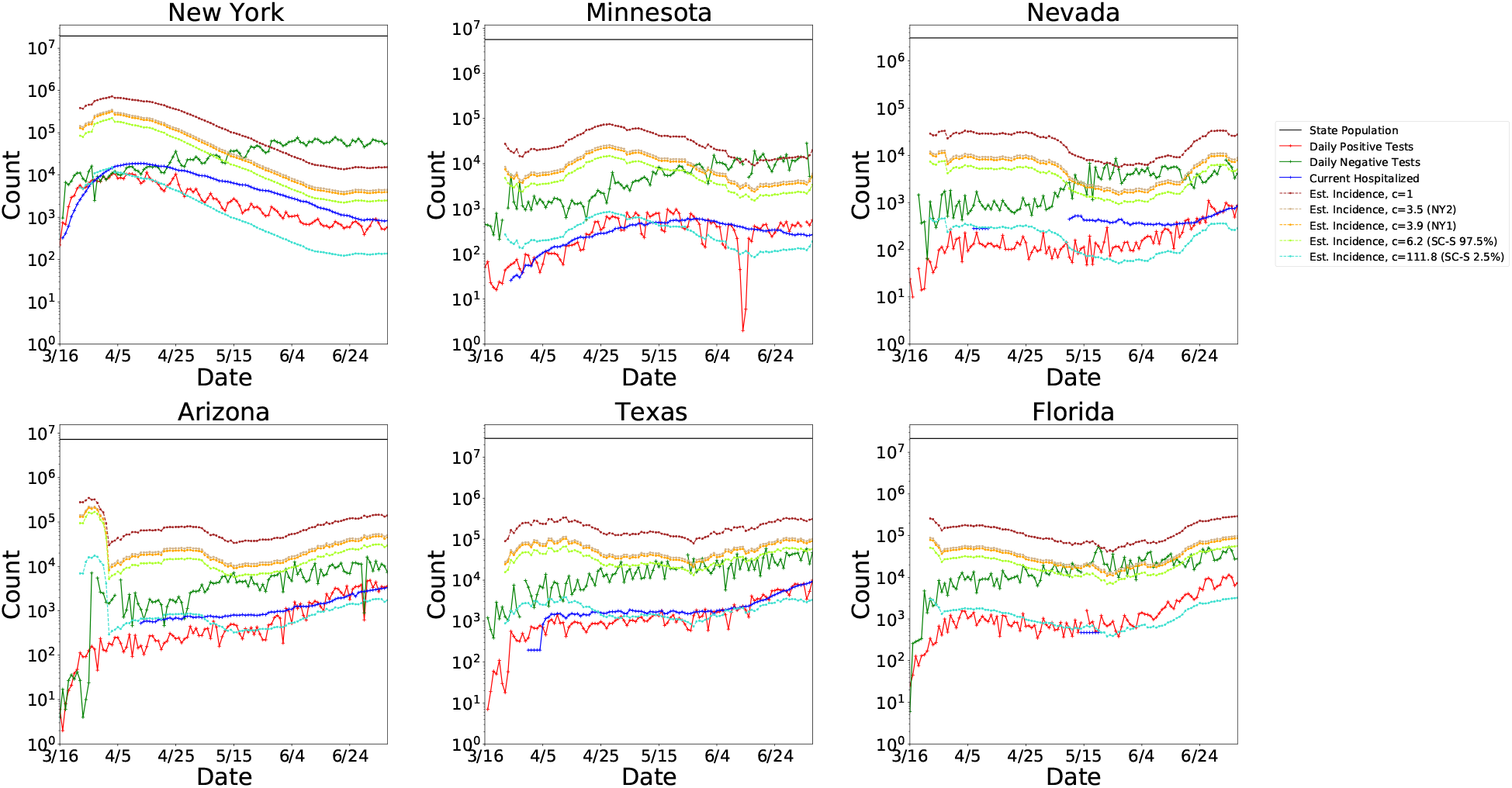
Representative results of applying the model Eq. (9) to estimate daily incidence from observed positive test counts, negative test counts, and the free parameter *c* (*t*), which represents the relative preference for testing infected individuals. For visual clarity, we smooth the estimated incidence curves by taking the weekly running average of daily counts. Shown here are results for six of fifty states, with estimated incidence based on several values of *c* (*t*) each implied by a different seroprevalence study.

**Figure 2:**
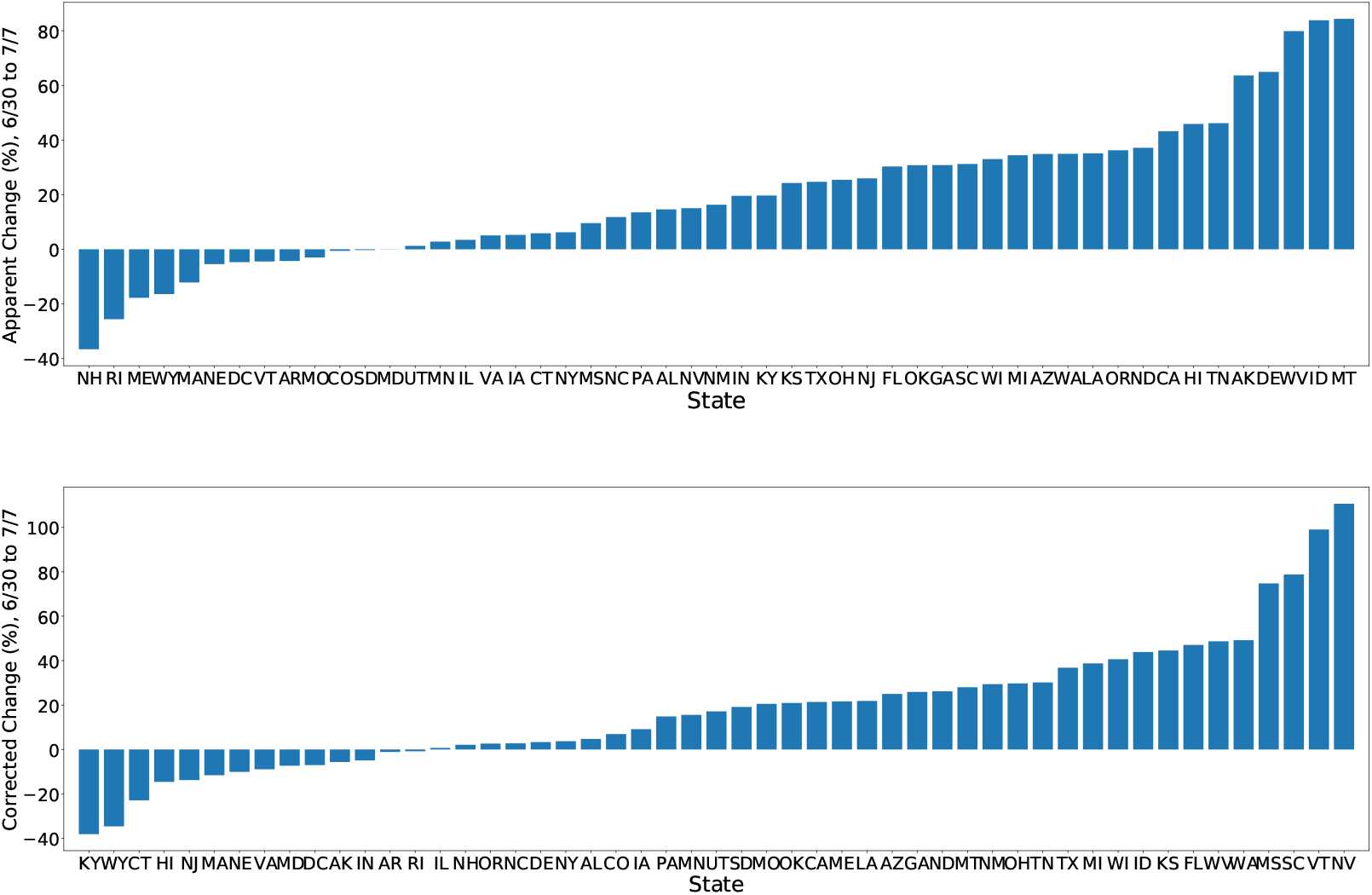
The unadjusted weekly spread of Covid-19 cases (top) generally outpaces the weekly spread adjusted for expanded testing (bottom). In some cases, such as NV, the adjusted spread could imply a larger problem than is obvious from the unadjusted increase in positive test counts.

As expected from mechanics of epidemic spread, both the number of positive tests and the estimated incidence were initially higher in the states with a higher population density than in the states with a lower population density (see Fig. 3 for a depiction of population densities). For example, the estimate of daily incidence in NY reached a peak in mid-April, while the estimates of daily incidence in AZ and TX have slowly increased over time. More recently, states such as MN, NV, AZ, TX, and FL have seen an uptick in the rate of positive test counts — according to our model, NV had the greatest level of epidemic spread.^2^

**Figure 3:**
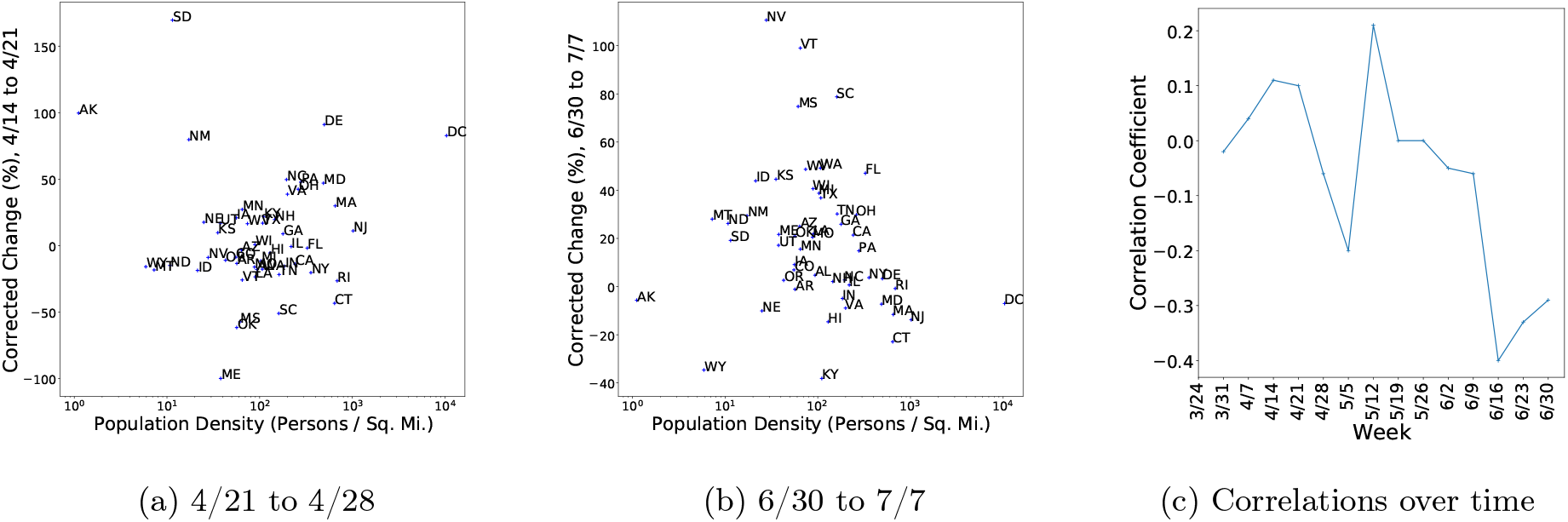
In the earlier months (left), there was a weak positive correlation between population density and epidemic spread. More recently (middle), there is a negative correlation between population density and epidemic spread. The Spearman correlation coefficient between population density and weekly change in estimated incidence (right) has gone from positive to negative through the course of the epidemic.

Finally, many of the curves of the estimated incidence grow at a slower rate than the rate of increase in the number of positive test results. This could happen for two reasons which cannot be distinguished without representative community testing: (1) either testing availability is expanding at a faster rate than the true epidemic is spreading, or (2) the true value of *c*(*t*) is decreasing over time. These explanations are not mutually exclusive, and in fact, (2) is likely to be a consequence of (1). We expect that the value of *c* (*t*) is decreasing over time because testing availability is increasing, and thus the curve of true incidence does not follow any one of the curves plotted for a fixed *c*(*t*). As the testing availability continues to increase and more serology studies further clarify the value of *c*(*t*), estimates of the incidence can be made with increasing accuracy.

### States Most at Risk for an Increase in Incidence Are Not Always the Same as States with the Most Positive Cases

Fig. 2 shows the weekly increase in positive SARS-CoV-2 tests (top) alongside the weekly change in our estimate of daily incidence (bottom). To calculate the corrected daily incidence, we compute Eq. 9 for each day. For both the uncorrected and the corrected incidences, we report the percentage change from the 7-day average ending on June 29, 2020 to the 7-day average ending on July 6, 2020. These results show that the majority of states have uncorrected levels of SARS-CoV-2 spread which are larger than the changes in the estimated incidence after correcting for increased testing.

In addition, the ranking of the states most at risk for increased SARS-CoV-2 spread changes after correcting for increased testing. The 5 states with largest percentage increase in positive test counts were MT, ID, WV, DE, and AK, while the 5 states with largest increase in our estimate were NV, VT, SC, MS, and WA.

We also examine the relationship between population density and the weekly change in the estimated incidence. As shown in Fig. 3, in the earlier months of the epidemic there was a weak positive correlation between population density and the weekly change in estimated incidence.^3^ However, in the summer months, we see that there is a negative correlation between these two features.

### Proportionally-Increasing Positive Counts are Indistinguishable from Expanding Testing Without Knowledge of Testing Protocols

A counter-intuitive statistical property which is made clear by this model is that an increasing number of positive tests does not necessarily increase the estimated incidence. Specifically, Eq. 5 shows that increasing the number of positive tests alongside a proportionate increase in the number of negative tests does not affect the estimated cumulative incidence. This represents the counter-intuitive property that if we observe only the counts of test results, an exponentially-spreading infection is statistically indistinguishable from exponentially-growing testing capacity unless we have extra information about the testing protocols. For instance, we can consider a infection which is fixed in some proportion of the full population (i.e. not spreading). In this hypothetical case, if testing were to grow at an exponential rate, then the counts of positive tests would grow at a similar exponential rate without the infection spreading at all. Thus, if we believe the incidence of the epidemic is truly increasing while the proportion of positive tests is remaining the same, then we must implicitly believe that the testing protocol is changing over time. Our parameter *c* (*t*) provides a way to quantify this change.

## Discussion

In this paper, we describe the development of a novel probabilistic model to estimate incidence of SARS-CoV-2 while adjusting for testing protocols. The model incorporates test results and a single, interpretable free parameter which was estimated from several external epidemiologic studies.

We used the results from our model to evaluate recent epidemic spread in each of the 50 states. We report substantial differences between unadjusted and adjusted incidence for selected states. These results can be used to help inform state policy-makers about the selection and implementation of public health policies and strategies during the Covid-19 pandemic. Our analysis also found that the state’s population density, which was previously positively correlated with increasing incidence, is more recently negatively correlated.

This study has several limitations. First, our results are dependent upon the quality of the external studies. However, we used several seroprevalence studies to provide a range of reasonable parameter values and minimize our dependence upon an individual study. Second, our model does not project future spread, such as could be done with a mechanistic model (e.g., SIR model; [1]). Third, our model uses test results, which are dependent on both test availability and usage, as well as test characteristics (e.g., sensitivity, specificity), for which we cannot correct.

This study also has important strengths which can be incorporated into future analyses of the incidence of SARS-CoV-2. First, we used several types of epidemiologic studies, as well as several seroprevalence studies, to estimate the free parameter, thus minimizing reliance upon a single external study or type. Second, this parameter can be re-estimated as results of new external studies become available. In particular, representative seroprevalence studies in localities with reported test counts will improve the estimation of the free parameter in our model. Third, our model has no other external inputs such as symptomatic rates which are difficult to estimate for a novel virus. Fourth, our model can be applied repeatedly and at various geographic levels to help inform public health decisions including re-opening of different localities.

In conclusion, we have developed a robust probabilistic model to estimate incidence and epidemic spread of SARS-CoV-2 infection. This model expands the set of methods and tools that public health officials can use to assess epidemic spread. Decision-makers can consider these estimates of incidence to inform public policy and avoid relying solely upon diagnostic test counts.

## Methods

We have *N* individuals in the population. At each time *t*, we observe the following random variables:

- *n*_*dp*_ (*t*), the number of individuals who test positive on day *t*
- *n*_*p*_ (*t*), the number of individuals who tested positive up to and including day *t*
- *n*_*dn*_ (*t*), the number of individuals who test negative on day *t*
- *n*_*n*_ (*t*), the number of individuals who tested negative up to and including day *t*

In addition, we have the unobserved latent variables:

- *z*^*i*^ (*t*), whether individual *i* has truly been infected
- *t*^*i*^ (*t*), whether individual *i* has been tested

Finally, let *T*_n_ (*t*) denote the probability that an uninfected individual will be tested, and *T*_p_ (*t*) denote the probability that an infected individual will be tested, i.e. *T*_*n*_(*t*) = 𝕡 (*t*^*i*^(*t*) = 1 | *z*^*i*^(*t*) = 0), and *T*_*p*_(*t*) = P(*t*^*i*^(*t*) =1 | *zi* (*t*) = 1). We then define

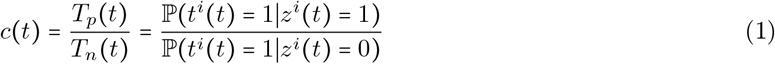

i.e. an infected individual is a factor of *c (t)* more likely to be tested than an uninfected individual.

### Inference of Cumulative Incidence

We would like to estimate the total number of individuals who have been infected by time *t*, 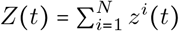 Given all *z*^*i*^(*t*), the expected number of positive tests is

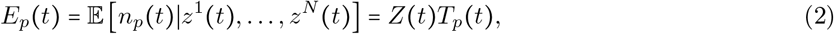

and the expected number of negative tests is

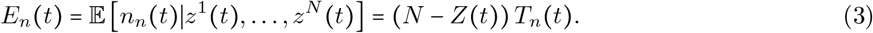

Therefore,

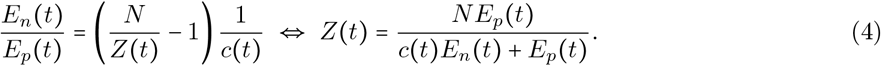

Based on this, we estimate *Z*(*t*) using

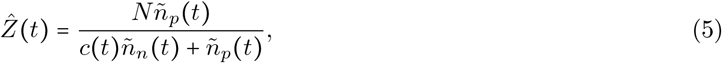

where *ñ*_*p*_(*t*) is the observed cumulative number of positive tests and *ñ*_*n*_(*t*) is the observed cumulative number of negative tests.

### Inference of Daily Incidence

To estimate the incidence for each day *t*, we must consider that only active infections will test positive under diagnostic testing. Let the length of an active infection be 14 days, and the day that an individual with an active infection is tested by uniformly distributed over the course of the infection. Then, the expected number of positive tests on day *t* is

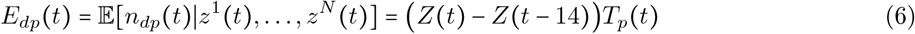

and the expected number of negative tests of day *t* is

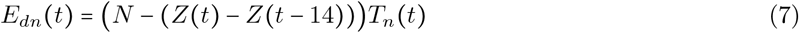

Therefore,

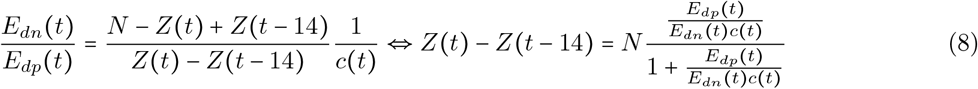

Based on this, we estimate the number of new infections on day *t* as

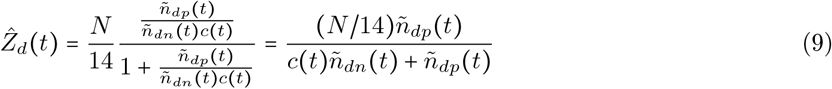

### Intuition

The central assumption underlying our model is that infected individuals are more likely than uninfected individuals to receive diagnostic tests. The parameter *c*(*t*) quantifies this bias toward testing SARS-CoV-2-infected patients: large values of *c*(*t*) indicate a large preference for testing SARS-CoV-2-infected patients, while a value of *c*(*t*) = 1 indicates no prejudice between testing SARS-CoV-2-infected and non-SARS-CoV-2-infected patients.

### Estimating *c* (*t*)

To estimate the free parameter *c* (*t*), we use seroprevalence as a measure of *Z* (*t*) and calculate the implied *c*(*t*). In contrast to the diagnostic tests, which have relied on patients seeking treatment for their disease, representative seroprevalence studies attempt to get an unbiased view of a region by randomly selecting participants and re-weighting demographic groups to reflect a broader population. Each study provides a view of *Z* (*t*) which implies a particular *c* (*t*) :

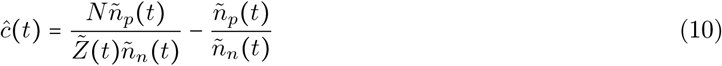

In Table 1, we show the value of *ĉ* (*t*) implied by the reported value of *Z* (*t*) in the following seroprevalence studies:

- **Santa Clara County, CA (SC) [2]:** Concerns have been raised ^4,5^ regarding improper propagation of uncertainty estimates in the original pre-print. In addition to the 95% confidence intervals reported in the original preprint (SC-B), we also report the 95% confidence interval calculated with the PyStan MCMC implementation ^6^ designed to correctly propagate uncertainties (SC-S).
- **New York, NY (NY1**^**7**^, **NY2**^**8**^): The state government has reported results of two seroprevalence studies. These results have been shared at press conferences and recorded in news articles; however, the full data is not currently available, so we calculate a point estimate of *ĉ* (*t*).
- **Boise, Idaho (ID) [3]**: An accurate antibody test used in Idaho’s “Crush the Curve” initiative in late April.

**Table 1:** Implications of seroprevalence reports.

| Study | Antibody Prevalence | Date | $\hat{c}(t)$ |
| --- | --- | --- | --- |
| SC-B | 0.0180–0.0570 | 4/4 | 1.74–5.74 |
| SC-S | 0.00094–0.0167 | 4/4 | 6.19–111.82 |
| NY1 | 0.139 | 4/21 | 3.92 |
| NY2 | 0.123 | 5/2 | 3.45 |
| ID | 0.0179 | 4/25 | 5.86 |

Each of these studies provide estimates of *Z*(*t*) but we do not view any of these values as the “correct” of *c*(*t*). We look forward to the release of more seroprevalence studies which can be matched with numbers values of positive and negative diagnostic tests to estimate more recent values of *c*(*t*).

### Related Work

A few other works have proposed estimating the cumulative incidence of SARS-CoV-2 from limited testing. One strategy is to estimate an “undertesting” ratio to understand how to correct the observed number of cases. Unfortunately, the undertesting ratio can be difficult to estimate for a novel disease because it requires knowledge of the distribution of outcomes from contact with the infectious agent. For example, [4] estimates the undertesting ratio by comparing case fatality ratios between localities, and arrive at an estimated undertesting ratio of approximately 0.5 for the United States. This ratio is likely to be drastically changing as testing availability changes, while the relative propensity-to-test is more consistent. As we learn more about the effects of SARS-CoV-2, we expect that estimates of undertesting ratios will be very helpful, but for this novel disease we prefer the relative propensity-to-test *c* (*t*) as an interpretable parameter.

Our results concord more with the approach of [5, 6] to use influenza-like illness (ILI) detection systems to estimate cumulative incidence of SARS-CoV-2. By using the pre-existing ILI detection systems to estimate the increase in ILI disease prevalence, [5] obtain estimates of the SARS-CoV-2 incidence which are 5-50 times higher than reported by diagnostic tests and [6] obtain estimates of the SARS-CoV-2 incidence which are also 5 times higher than reported by diagnostic estimates. This magnitude of undercounting has also been reported by more recent analyses of city-level seroprevalence [7]. Our approach using the relative propensity-to-test (*c* (*t*)) suggests that this undercounting magnitude may be realistic.

## Data Availability

Test count data is collected by the Covid Tracking Project9. For each state, we set the population *N* according to the 2019 state population estimates projected from the 2010 US Census^10^.

## Data Availability

Test count data is collected by the Covid Tracking Project. Python code to reproduce the analysis is available at the following link.

https://github.com/blengerich/Covid19-LatentCases

## Footnotes

1 https://github.com/blengerich/Covid19-latentcases

2 The number of positive tests are unreliable in MN in early June, so it is difficult to infer the extent of the epidemic spread in that state.

3 State-level counts of positive and negative test results were not available before late March, so the initial outbreak of the epidemic is not captured in these figures.

4 https://statmodeling.stat.columbia.edu/2020/04/19/fatal-flaws-in-stanford-study-of-coronavirus-prevalence

5 https://twitter.com/AlanMCole/status/1251876582211432448

6 https://statmodeling.stat.columbia.edu/2020/04/19/fatal-flaws-in-stanford-study-of-coronavirus-prevalence/#comment-1305950

7 https://www.nbcnewyork.com/news/local/new-york-virus-deaths-top-15k-cuomo-expected-to-detail-plan-to-fight-nursing-home-outbreaks/2386556/

8 https://twitter.com/NYGovCuomo/status/1256602494165225472

9 https://covidtracking.com/data/

10 https://www.census.gov/newsroom/press-kits/2019/national-state-estimates.html

## References

[1] Ottar N Bjørnstad, Katriona Shea, Martin Krzywinski, and Naomi Altman. Modeling infectious epidemics. Nature methods, 2020.

[2] Eran Bendavid, Bianca Mulaney, Neeraj Sood, Soleil Shah, Emilia Ling, Rebecca Bromley-Dulfano, Cara Lai, Zoe Weissberg, Rodrigo Saavedra, James Tedrow, et al. Covid-19 antibody seroprevalence in santa clara county, california. medRxiv, 2020.

[3] Andrew Bryan, Gregory Pepper, Mark H. Wener, Susan L Fink, Chihiro Morishima, Anu Chaudhary, Keith Jerome, Patrick C Mathias, and Alex Greninger. Performance characteristics of the abbott architect sars-cov-2 igg assay and seroprevalence testing in idaho. medRxiv, 2020.

[4] Timothy W Russell, Joel Hellewell, Sam Abbott, CI Jarvis, K van Zandvoort, CMMID nCov working group, S Flasche, AJ Kucharski, et al. Using a delay-adjusted case fatality ratio to estimate under-reporting. Centre for Mathematical Modeling of Infectious Diseases Repository, 2020.

[5] Fred S Lu, Andre T Nguyen, Nicholas B Link, Marc Lipsitch, and Mauricio Santillana. Estimating the early outbreak cumulative incidence of covid-19 in the united states: Three complementary approaches. medRxiv, 2020.

[6] Justin D. Silverman, Nathaniel Hupert, and Alex D. Washburne. Using influenza surveillance networks to estimate state-specific prevalence of sars-cov-2 in the united states. Science Translational Medicine, 2020.

[7] Fiona P. Havers, Carrie Reed, Travis W. Lim, Joel M. Montgomery, John D. Klena, Aron J. Hall, Alicia M. Fry, Deborah L. Cannon, Cheng-Feng Chiang, Aridth Gibbons, Inna Krapiunaya, Maria Morales-Betoulle, Katherine Roguski, Mohammed Rasheed, Brandi Freeman, Sandra Lester, Lisa Mills, Darin S. Carroll, S. Michelle Owen, Jeffrey A. Johnson, Vera A. Semenova,, Jarad Schiffer, and Natalie P. Thornburg. Seroprevalence of antibodies to sars-cov-2 in six sites in the united states, march 23-may 3, 2020. medRxiv, 2020.

